# Dengue nowcasting in Brazil by combining official surveillance data and Google Trends information

**DOI:** 10.1101/2024.09.02.24312934

**Authors:** Yang Xiao, Guilherme Soares, Leonardo Bastos, Rafael Izbicki, Paula Moraga

## Abstract

Dengue is a mosquito-borne viral disease that poses significant public health challenges in tropical and sub-tropical regions worldwide. Surveillance systems are essential for dengue prevention and control. However, traditional systems often rely on delayed data, limiting their effectiveness. To address this, nowcasting methods are needed to estimate underreported cases, enabling more timely decision-making. This study evaluates the value of using Google Trends indices of dengue-related keywords to complement official dengue data for nowcasting dengue in Brazil, a country frequently affected by this disease. We compare various nowcasting approaches that incorporate autoregressive features from official dengue cases, Google Trends data, and a combination of both, using a naive approach as a baseline. The performance of these methods is evaluated by nowcasting weekly dengue cases from March to June 2024 across Brazilian states. Error measures and 95% coverage probabilities reveal that models incorporating Google Trends data enhance the accuracy of weekly nowcasts across states and offer valuable insights into dengue activity levels. To support real-time decision-making, we also present Dengue Tracker, a website that displays weekly dengue nowcasts and trends to inform both decision-makers and the public, improving situational awareness of dengue activity. In conclusion, the study demonstrates the value of digital data sources in enhancing dengue nowcasting, and emphasizes the value of integrating alternative data streams into traditional surveillance systems for better-informed decision-making.

**Author summary:** Dengue is a mosquito-borne viral disease that poses significant public health challenges in tropical and sub-tropical regions worldwide. Surveillance systems are crucial for dengue prevention and control. Unfortunately, traditional systems often rely on delayed data, limiting their effectiveness. To address this, nowcasting methods are needed to estimate underreported cases, enabling more timely decision-making. This study evaluates how Google Trends indices of dengue-related keywords can complement official dengue data to improve nowcasting of dengue in Brazil, a country frequently affected by this disease. We compare the performance of various nowcasting approaches that incorporate Google Trends data with other approaches that rely solely on official reported cases data, assessing their accuracy and uncertainty in nowcasting weekly dengue cases from March to June 2024 across Brazilian states. To support real-time decision-making, we also present Dengue Tracker, a website that displays weekly dengue nowcasts offering valuable insights into dengue activity levels. The study demonstrates the potential of digital data sources in enhancing traditional surveillance systems for better-informed decision-making.

## Introduction

Dengue is a viral disease transmitted by mosquitoes of the *Aedes* genus such as *Aedes aegypti* and *Aedes albopictus*, which poses a significant global health threat, particularly in tropical and subtropical regions like Latin America, Southeast Asia, the Pacific Islands, parts of the Middle East, and Africa (1). The disease ranges in severity from mild symptoms such as high fever, severe headache, retro-orbital pain, joint and muscle pain, and rashes, to severe forms like dengue hemorrhagic fever. This severe condition can lead to critical complications such as bleeding, plasma leakage, and organ failure, significantly increasing the risk of mortality (2).

Dengue transmission occurs through mosquito bites, with peak activity early in the morning and before dusk. The disease is highly contagious and can rapidly spread among populations, particularly in densely populated urban areas where mosquitoes breed in stagnant water. Several factors contribute to the prevalence of dengue, including climate, urbanization, and socio-economic disparities (3). Additionally, global warming and increased international travel have expanded the geographic range of dengue (4).

Annually, there are approximately 400 million infections, with 100 million cases exhibiting clinical symptoms ranging from mild to severe (2; 5). In 2024, Brazil is facing a severe dengue outbreak, with 9.48 million suspected cases and 5.32 million confirmed cases as of August (6). This surge has made Brazil the most affected country in the Americas. The increased incidence is attributed to factors like early transmission seasons, climate change, and the presence of all four dengue serotypes.

Challenges in dengue prevention and control include the absence of specific antiviral treatments and the complexity of developing vaccines for the virus’s four serotypes (7). Moreover, dengue symptoms can resemble those of other diseases such as chikungunya and Zika leading to misdiagnosis and complicating effective control measures. Dengue prevention efforts focus on reducing mosquito populations and avoiding bites through community engagement, personal protection, and environmental measures.

The rapid transmission and extensive impact of dengue underscore the importance of research and the ability to predict its outbreaks. Surveillance systems play a crucial role for guiding strategies for prevention and control, but traditional surveillance systems often rely on delayed or incomplete data due to underreporting, healthcare infrastructure limitations, and time lags in laboratory testing. For example, in Brazil, a suspected dengue case is required by law to be reported to the government authorities. The notification is made by authorized health professionals in Notifiable Diseases Information System (SINAN) (8). The notification data is then gathered in Brazilian Ministry of Health and the most up-to-date information is used by the Epidemiological Surveillance teams to understand current status of dengue cases through the country. However, according to (9), less than 50% of dengue cases are reported within the first week, no more than 75% are reported within four weeks, and fewer than 90% are reported within nine weeks. This latency necessitates nowcasting methods to estimate occurred-but-not-yet-reported disease cases for real-time decision-making.

Several nowcasting techniques have been developed in different settings. For example, (10) employed reverse-time discrete hazard functions and maximum likelihood estimation to handle reporting delays and nowcast AIDS cases in Canada. A Bayesian hierarchical model was proposed by (11) to improve prediction and management of a Shiga toxin producing Escherichia coli that caused a major outbreak in Germany in 2011. The model combined a survival regression model for the delay distribution, and a quadratic spline for the epidemic curve, utilizing the generalized Dirichlet distribution for flexibility in handling uncertainty. (12) proposed a Bayesian hierarchical model that jointly estimates the expected number of deaths, and the reporting delay distribution to nowcast COVID-19 fatalities in Sweden. This model offers enhanced predictive performance and flexibility by incorporating leading indicators such as the number of reported cases and COVID-19 associated ICU admissions.

A nowcasting by Bayesian smoothing approach capable of producing nowcasts in multiple disease settings was developed by (13). This approach learns the reporting delay distribution and the time evolution of the epidemic to produce nowcasts in both stable and time-varying case reporting settings. The approach was tested on dengue in Puerto Rico and influenza-like illness (ILI) in the United States. (14) presented a framework for addressing reporting delays in malaria surveillance in Guyana. The method combines a data imputation model and network models to refine case estimates using historical data, neighboring region data, and precipitation levels. (15) introduced a Bayesian framework with sliding windows for dengue surveillance in Bangkok, Thailand, addressing reporting delays by accounting for spatial and temporal variations. A Bayesian hierarchical model for dengue nowcasting in Brazil was developed by (9). This approach uses a Negative Binomial distribution for the reported cases with mean explained by spatial, temporal, and delay information, offering a robust correction to the reported cases. This model is used by the InfoDengue system to nowcast dengue in Brazilian municipalities (16).

The implementation of these methods are complex and require extensive data on the historical weekly reported cases for the disease under consideration. In recent years, methods utilizing digital data, such as Google Trends indices and Twitter (now X) information on disease-related keywords, have shown significant effectiveness in understanding and predicting disease activity levels. These methods leverage real-time search query data to enhance the accuracy of traditional models, allowing for more timely and reliable public health responses.

For example, (17) utilized used search query logs and modeling techniques such as Elastic Net regularized regression and Gaussian Process regression to nowcast influenza-like illness in the USA. (18) used an ARIMA model augmented with Google Flu Trends data for nowcasting influenza outbreaks in the USA. They showed the incorporation of real-time search query data improves prediction accuracy compared to a model that uses only case data.

A Hidden Markov Model combining cases and Google Trends information for disease prediction was proposed by (19). The model incorporated an autoregressive component describing case counts and a linear covariate representing Google Trends. The model was applied to predict dengue in Brazil, Mexico, Thailand, Singapore, and Taiwan, as well as influenza-like illness in the USA (20).

In (21), authors utilized Baidu search query data, which is similar to Google Trends, to nowcast hand, foot, and mouth disease across China. They utilized a meta-learning framework to dynamically select among predictive models including Principal Component Analysis, LASSO, Ridge Regression, and ARIMA. They showed the inclusion of Baidu Index data enhances prediction accuracy by providing real-time public interest metrics correlated with hand, foot, and mouth cases.

(22) used Twitter data to monitor dengue in Brazil. First, they analyzed tweet sentiments to filter tweets indicative of actual cases, and found a high correlation of the number of dengue-related tweets with official dengue data. Then, they constructed a regression model for predicting the number of dengue cases using the proportion of dengue-related tweets, and used it to develop a monitoring system that generated weekly heatmaps of dengue across cities in Brazil.

We are not aware of any approach that utilizes Google Trends data in Brazil. In fact, the only existing dengue tracking system in Brazil is InfoDengue, which provides weekly case reports of dengue and other arboviruses across Brazilian municipalities, along with nowcasts generated by a Bayesian hierarchical model (16). Therefore, it is crucial to investigate alternative methods.

To this end, in this paper we assess the value of Google Trends to nowcast weekly dengue cases in Brazilian states by fitting models that integrate reported dengue cases in Brazilian states provided by InfoDengue https://info.dengue.mat.br/, and Google Trends indices of dengue-related keywords at the state level in Brazil which can be obtained from https://trends.google.com/trends/. Then, we compare nowcasts produced by models that utilize only reported dengue cases, only Google Trends data, and a combination of both, with the model provided by InfoDengue (16). As a baseline, we also use a naive approach where nowcasts are considered as the reported cases the previous week. Our aim is to evaluate the value of real-time Google Trends data in producing accurate nowcasts using simple models that do not rely on incomplete recent case data, and to determine whether these nowcasts can compete with more complex and time-consuming models.

Additionally, recognizing the importance of timely and accurate data for dengue surveillance, we developed the Dengue Tracker website (https://diseasesurveillance.github.io/dengue-tracker/). This site is updated with weekly nowcasts for each Brazilian state, and presents information through interactive maps and time trend plots to inform decision-makers and the public about dengue activity levels in real-time.

The rest of the paper is organized as follows. First, we detail the study region, data sources, nowcasting methods, and performance assessment measures. Next, results are presented through explanatory tables and figures, highlighting error and uncertainty measures across states. Then, we describe the Dengue Tracker website developed to support real-time decision-making. Finally, the paper concludes with a discussion of the findings and future work.

## Materials and methods

### Study region

Brazil, the largest country in South America and the fifth-largest in the world, spans a vast geographical area characterized by diverse climates and ecosystems (23). Its territory stretches from the equatorial Amazon basin in the north to the temperate regions in the south. This wide latitudinal range encompasses various climatic zones, including tropical, subtropical, and temperate, which significantly influence the epidemiology of vector-borne diseases like dengue fever. Brazil is divided into 26 states and one federal district, each with unique geographical and socio-economic characteristics that impact dengue transmission and control (Figure 1).

**Fig 1.**
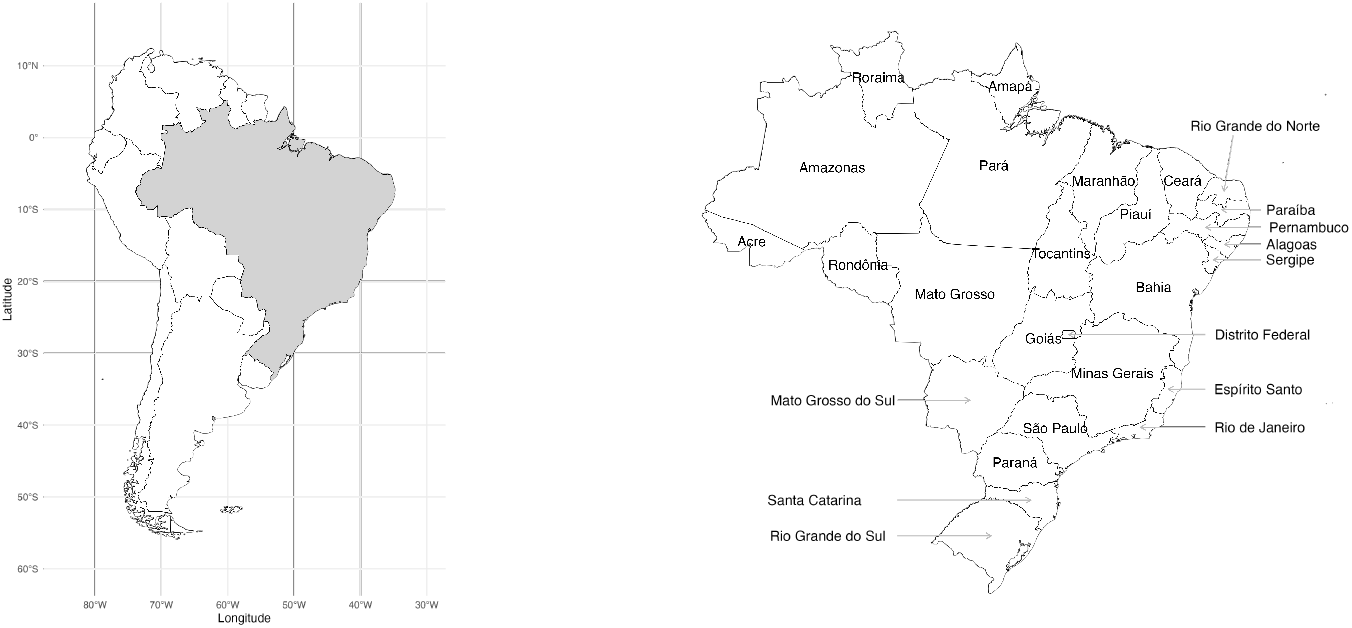
Map of South America with Brazil highlighted (left) and map of 26 states and the federal district of Brazil (right).

Several factors significantly contribute to the proliferation of dengue outbreaks in Brazil (24). Climate change, characterized by increased temperatures, has diminished geographical barriers to dengue transmission, particularly in southern Brazil, by reducing the seasonal cold periods that typically inhibit mosquito propagation. In the Amazon region, climatic changes have made previously protected areas more susceptible to dengue outbreaks.

Rapid urbanization has led to high population densities in large cities, fostering environments conducive to mosquito breeding due to inadequate infrastructure, such as insufficient piped water and waste management systems (25). This results in standing water containers and uncollected garbage, serving as breeding grounds for mosquitoes. Furthermore, high connectivity within Brazil’s urban network exacerbates the risk of disease spread. Major regional hub cities with frequent transportation and logistical connections facilitate faster virus transmission, while enhanced connectivity between urban and rural areas contributes to the spread of the virus from cities to the countryside (24).

Figure 2 illustrates the seasonal pattern of monthly dengue incidence rates in Brazilian states from January 2010 to July 2024, aggregated at the state level, with data sourced from InfoDengue (26). The figure reveals a seasonal pattern, with dengue outbreaks typically occurring from January to May. In certain regions, such as Acre, Rondônia, Mato Grosso, and Goiás in southwestern Brazil, outbreaks may even commence in November or December. The spread of the disease is generally minimal during the winter months. Moreover, it is observed that dengue outbreaks tend to be more severe every three to four years. This periodicity could be influenced by El Niño, which significantly impacts weather patterns in certain areas of Brazil (27). Notably, the states of Acre, Espírito Santo, Goiás, Mato Grosso do Sul, Paraná, Rio Grande do Norte, São Paulo, and Tocantins experience a more substantial impact from dengue, as indicated by consistently higher incidence rates.

**Fig 2.**
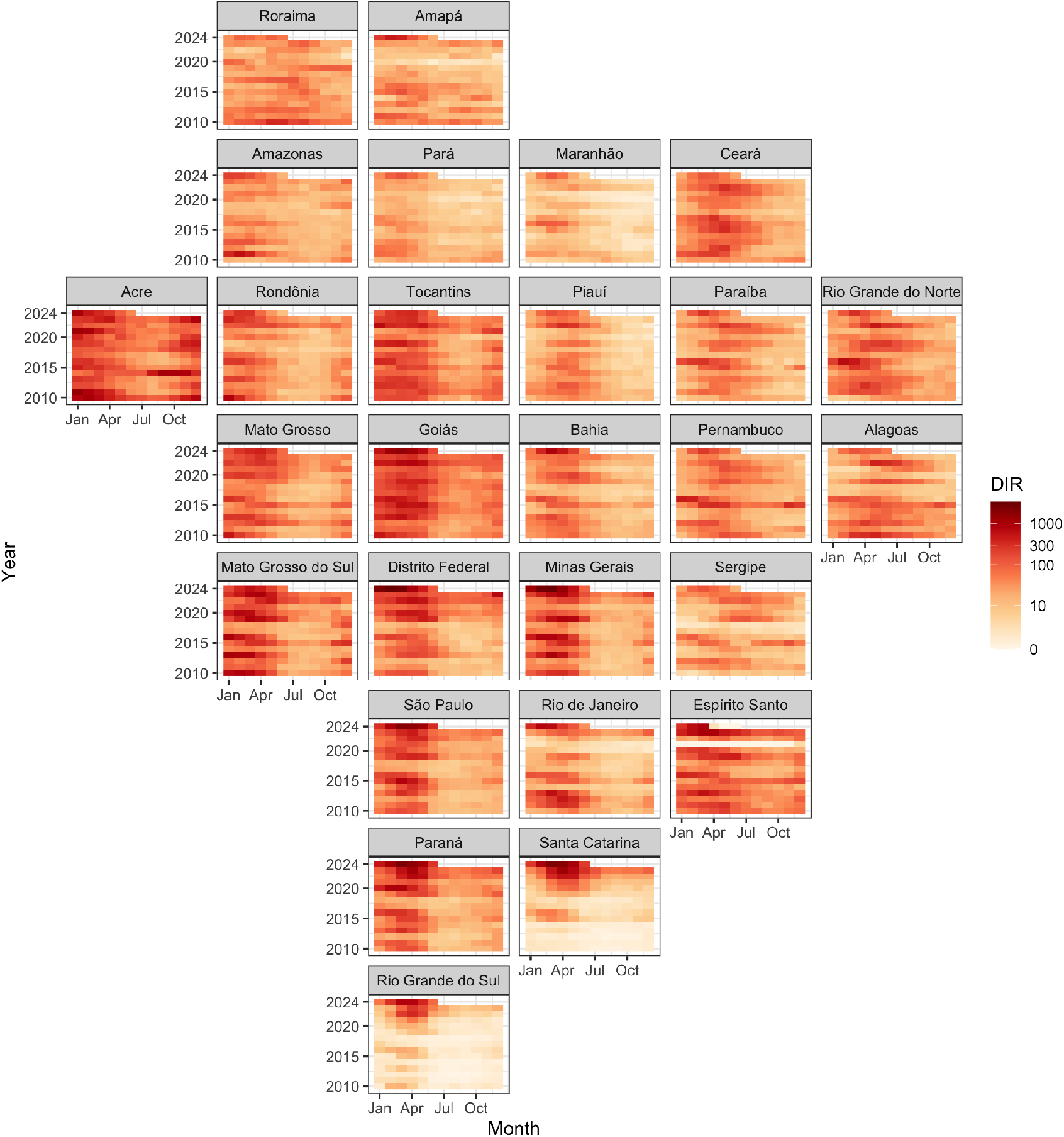
Dengue incidence rate (cases per 100k people) on a *log*_10_ scale in Brazilian states from January 2010 to July 2024.

### Dengue cases data

The InfoDengue system (28) provides comprehensive data on dengue and other arboviruses across Brazil, offering detailed insights into regional variations, trends, and severity of outbreaks. Accessible at https://info.dengue.mat.br/, the platform aggregates data provided from SINAN to present dengue cases by epidemiological week and municipality.

Despite being a reliable source of information, the cases shown in InfoDengue suffer from reporting delays. As mentioned in (9), although in principle dengue is meant to be reported within seven days, in practice no more than 90% of the cases are reported within 9 weeks. Therefore, we consider as the actual number of dengue cases the number of cases reported in the system after 10 weeks. The difference between the provisional and final dengue cases represents the delay that needs to be accounted for in the nowcasting models. In addition, InfoDengue provides nowcasts and comprehensive visualizations into the geographic and temporal distribution of dengue, making it an essential tool for timely decision-making and proactive response to emerging outbreaks.

### Google Trends data

Google Trends (https://trends.google.com/trends/) is a tool that provides anonymized and aggregated insights into global search behaviors, ensuring user privacy by anonymizing individual searches and consolidating them into high-level trends. This data is valuable for identifying emerging topics and seasonal trends. The Google Trends index for a specific keyword at a given time ranges from 0 to 100, calculated by dividing the number of searches for that keyword by the total number of searches in a specific region and timeframe, enabling fair comparisons between search terms, locations, and periods. This standardization eliminates disparities due to population size or total search volume. However, it may also create misleading representations, as a term could appear less popular in larger regions due to a dilution effect. Moreover, aggregation might obscure specific nuances or emerging trends within smaller subgroups, potentially limiting the depth of analysis (29).

Here, we utilize Google Trends data to understand dengue search behavior patterns that could complement official dengue data. To select the keywords for the Google Trend indices to include in the models, we calculated the Pearson correlation between country-level aggregated dengue cases and the Google Trends index for various dengue-related keywords identified in (30). We used data from January 1, 2013, to December 31, 2023, with correlations computed using monthly resolution data, as Google Trends data cannot be obtained at a finer temporal resolution for periods exceeding five years and two months.

Figure 3 illustrates the correlation between the number of dengue cases and the Google Trends index corresponding to each of the keywords considered. We observe the highest correlation of dengue cases and Google Trends index corresponding to keywords “sintomas dengue” “dengue” and “sintomas de dengue” with correlations of 0.93, 0.90, and 0.89, respectively. The intercorrelations among these keywords were also significant, with the highest correlation being 0.97 between “sintomas dengue” and “dengue.”

**Fig 3.**
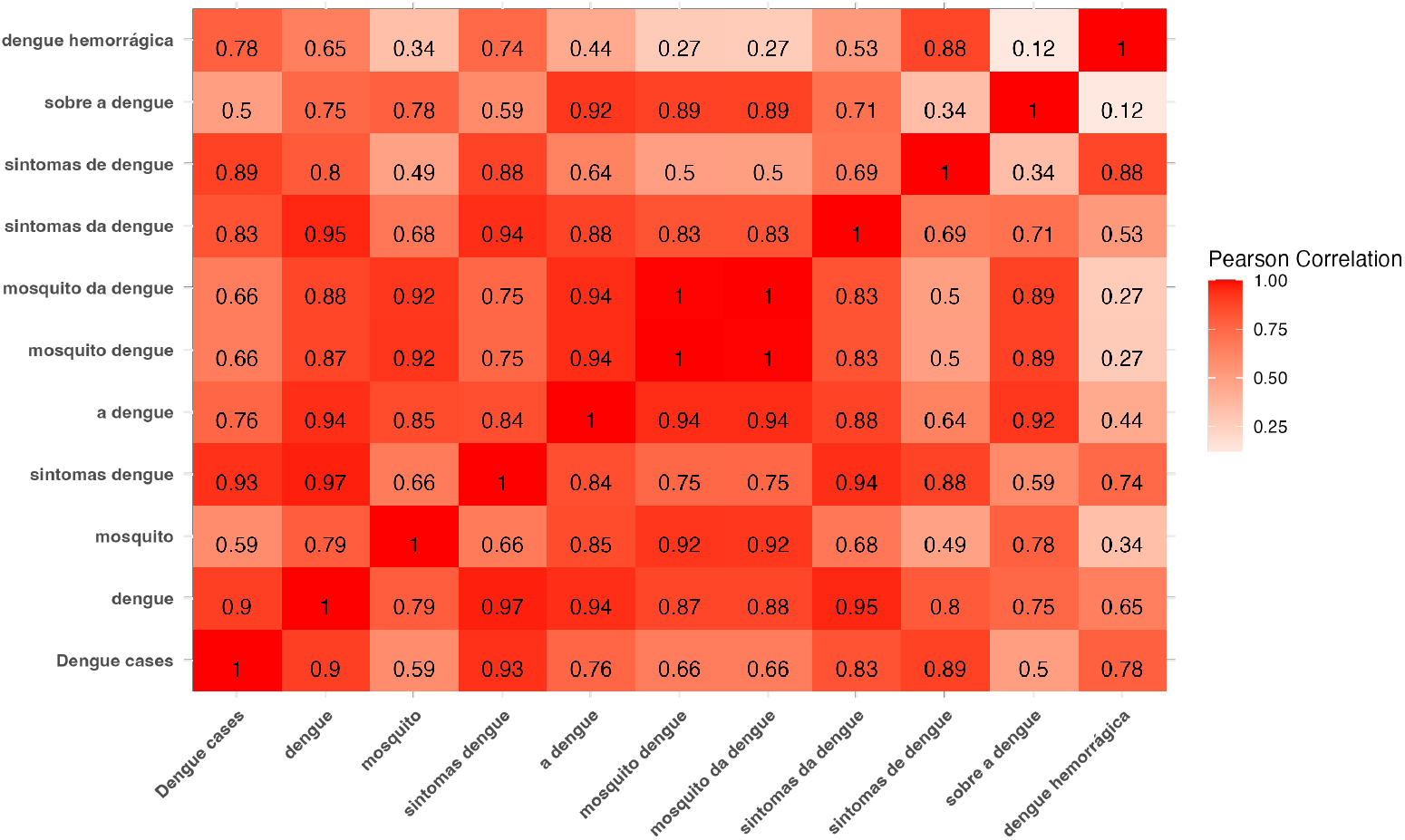
Correlation among dengue cases and Google Trends indices for several dengue-related keywords in Brazil.

In addition, to assess the current situation, we used Google Keyword Planner https://ads.google.com/intl/en/home/tools/keyword-planner/ to find keywords related to “dengue” in Brazil from January to April 2024. By providing the keyword dengue, the tool showed high search interest in “dengue” and “sintomas de dengue.” Consequently, we decided to incorporate Google Trends indices for the keywords “sintomas dengue” and “dengue” into our models.

### Nowcasting methods

We evaluate the performance of several approaches for nowcasting the weekly number of dengue cases in each Brazilian state. Specifically, we use five approaches: 1) a model that uses only official dengue case data; 2) a model that uses only Google Trends data; 3) a model that integrates both official dengue case data and Google Trends indices; 4) a Bayesian nowcasting approach implemented in the InfoDengue system; and 5) a naive approach that predicts cases based on the previous week’s data.

Models are evaluated using a moving window strategy, where each model is trained on a fixed-size window of historical data. Nowcasts are then generated for the last week of the window based on this training. The window advances by one week iteratively, producing a sequence of nowcasts over time that will be compared with the actual number of cases using several error and uncertainty measures.

In this study, we began recording the weekly number of dengue cases from InfoDengue on epidemiological week 10 of 2024 (March 3). Due to reporting delays, the initially reported number of cases is significantly lower than the actual number. Each week, InfoDengue updates the case numbers, continuing this process for up to approximately 10 weeks (9). Therefore, we assume the actual number of cases is accurately reported after a ten-week delay.

We started by producing a nowcast for week 10 of 2024 (March 3, 2024), using models trained on data from the past three years, from epidemiological week 6 in 2021 to epidemiological week 6 in 2024 (February 7, 2021, to February 4, 2024), 156 weeks in total. The models were trained using data that excluded the four most recent weeks, and provided a nowcast for the current number of dengue cases (four weeks ahead). We decided to exclude the most recent four weeks of training data to balance maintaining recent information with discarding incomplete data. During this period, the reported dengue cases do not accurately reflect the actual numbers, with less than 75% of cases being reported within four weeks (9).

This procedure is repeated by moving the window forward by one week for fourteen weeks, obtaining nowcasts for epidemiological weeks 10 to 23 of 2024 (March 3 to June 2, 2024). By computing error metrics and uncertainty intervals over multiple windows, this approach is particularly useful for validating models in dynamic and seasonal contexts, providing robust insights into nowcasts accuracy.

Let *c*_*t*_ represent the actual number of dengue cases at week *t*, and let *y*_*t*_ represent the official number of dengue cases reported at week *t*. As previously discussed, *y*_*t*_ is lower than *c*_*t*_ due to reporting delays, and we are interested in obtaining a nowcast *ĉ*_*t*_. Here, we assume the actual number of cases *c*_*t*_ is the number of dengue cases reported in InfoDengue 10 weeks after *t*. In addition, let *x*_*n,t*_ be the Google Trends index for the *n*th keyword at week *t*.

The nowcasting approaches considered include DC, GT, and DCGT, which use information only from dengue cases, only from Google Trends, and a combination of both datasets, respectively. These models use historical reported dengue cases *y*_*t*_ excluding the most recent weeks to produce nowcasts *ĉ*_*t*_. That is, the models are trained using only data for which we expect *y*_*t*_ ≈ *c*_*t*_. In addition, a Bayesian nowcasting model and a naive approach are also considered. The descriptions of the five nowcasting approaches are as follows.

### DCGT

The DCGT model uses SARIMA with eXogenous factors (SARIMAX) (31) to combine dengue cases (time series) with Google Trends indices (exogenous factor). The SARIMAX model is represented by a set of parameters equal to (*p, d, q*) *×* (*P, D, Q, S*), where *p* represents the order of auto-regression, *q* is the the order of moving-average, and *d* symbolizes differencing by which non-stationary time series are transformed into stationary time series. *P, D, Q, S* represent the combination of the Seasonal ARIMA (SARIMA) component. The exogenous part *x* is a variable outside the model, imposed on it but not influenced by it.

Here, the time series *y*_*t*_ can be written as the mathematical formulation of a SARIMA (*p, d, q*) *×* (*P, D, Q, S*) model with exogenous variable as follows:

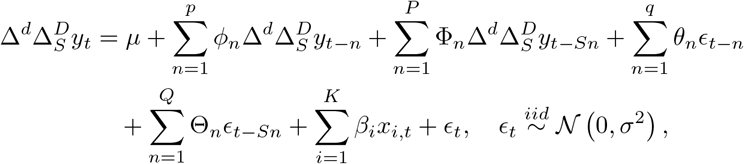

where 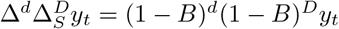 and *B* is the back-shift operator.

### DC

The DC model employs a Seasonal Autoregressive Integrated Moving-Average (SARIMA) model using dengue case data. It is the same as the DCGT model but removing the exogenous regressor.

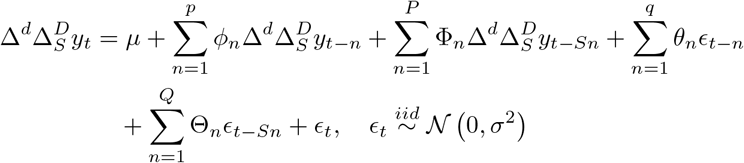

### GT

The GT model uses a linear model using an intercept and the Google Trends indices for the keywords ”dengue” and “sintomas dengue” as covariates.

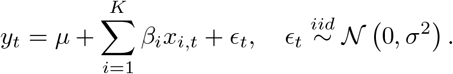

### InfoDengue

InfoDengue provides nowcasts using a Bayesian hierarchical model where the observed number of events *n*_*t,d,s*_ at time *t* reported after *d* time units in spatial location *s* is assumed to follow a Negative Binomial distribution with mean *λ*_*t,d,s*_ and dispersion *ϕ* (9). Specifically,

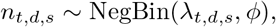

with mean E[*n*_*t,d,s*_] = *λ*_*t,d,s*_ and variance Var[*n*_*t,d,s*_] = *λ*_*t,d,s*_(1 + *λ*_*t,d,s*_*/ϕ*). To capture the temporal and spatial variability of *n*_*t,d,s*_, the mean is expressed as

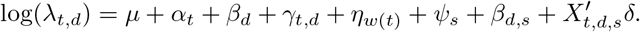

Here, *μ* represents the overall mean on the log scale, and 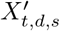 is a matrix of temporal, delay-related, and spatially varying covariates with associated parameter vector *δ. α*_*t*_ and *β*_*d*_ capture time and delay structure means, respectively, modeled as first-order random walks. The model also includes random effects *γ*_*t,d*_ to capture the interaction between time and delay, and a seasonal component *η*_*w*(*t*)_. Finally, *ψ*_*s*_ represents spatial variability, and *β*_*d,s*_ captures how the delay structure varies across different spatial locations.

### Naive

The naive approach uses the number of cases reported in the previous week as the nowcast of week *t*:

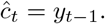

### Accuracy and uncertainty metrics

We assessed the performance of each nowcasting approach using several error measures. In addition, we computed the 95% coverage probabilities, which represent the proportion of times actual cases were covered by the 95% uncertainty intervals, as well as the average width of these intervals. Let *ĉ*_*t*_ and *c*_*t*_ represent the predicted and actual number of dengue cases, respectively, at time *t*, where *t* = 1, …, *n*. The error measures computed include the Root Mean Squared Error (RMSE), which measures the square root of the average squared differences between the predicted and actual values as

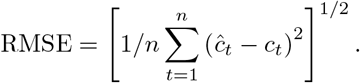

The RMSE emphasizes large errors due to the squaring process, making it sensitive to outliers. We also calculated the Mean Absolute Error (MAE) as the average absolute differences between predicted and actual values:

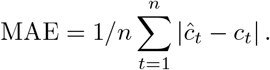

The MAE provides a straightforward interpretation of the average error magnitude which is less sensitive to outliers compared to RMSE. In addition, we computed the Root Mean Squared Percentage Error (RMSPE) as the square root of the average squared percentage errors:

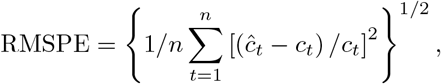

and the Mean Absolute Percentage Error (MAPE) as the average absolute percentage differences between predicted and actual values:

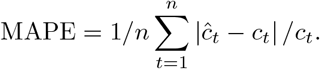

These measures are useful when the relative error is more meaningful than the absolute error, highlighting proportional discrepancies.

### Implementation

All analyses were performed using the statistical software R (32). Nowcasts and 95% confidence intervals for the DCGT and DC models were obtained using the function auto.arima() from the forecast package using the default maximal orders (33). Results for the GT model were obtained with the linear model function lm() from R. For reproducibility purposes, data and code to apply these methods are provided in the GitHub repository https://github.com/diseasesurveillance/dengue-tracker/tree/main/paper.

## Results

This section presents the error and uncertainty measures obtained for each method across all states. Results are not shown for the Espírito Santo as this state stopped reporting dengue cases to the federal governement since epidemiology week 16 of 2024 (April 14), resulting in missing case counts from that date onwards. Furthermore, the data for epidemiology week 24 of 2024 (June 9) was not uploaded by InfoDengue, so this week’s comparison was skipped in our analysis. For comparisons requiring this week’s data as the “true value”, the data from epidemiology week 25 of 2024 (June 16) was used.

Tables 1 to 4 present the error measures obtained for each state and nowcasting approach, where red indicates the best performance (smallest error), and blue represents the second-best performance (second smallest error) model. Figure 4 displays boxplots of the differences between the nowcasts and the true number of cases across states. In this figure, models are sorted by the mean of the differences being closest to zero, corresponding to the MAE. Maps with the best nowcasting models across states according to each error metric are shown in Figure 5.

**Table 1.** RMSE obtained for each state and nowcasting approach. Red and blue represent the best and the second best performances respectively (the lowest and the second lowest error).

|  | DCGT | DC | GT | InfoDengue | Naive |
| --- | --- | --- | --- | --- | --- |
| Acre | 238.58 | 236.52 | 96.46 | 252.41 | 261.92 |
| Alagoas | 384.5 | 416.68 | 681.75 | 299.43 | 508.08 |
| Amapá | 307.5 | 275.27 | 125.18 | 143.42 | 315.95 |
| Amazonas | 143.9 | 424.48 | 108.62 | 317.05 | 332.88 |
| Bahia | 10638.59 | 11927.25 | 6839.64 | 16064.65 | 8836.69 |
| Ceará | 856.19 | 969.44 | 2250.78 | 674.55 | 1021.84 |
| Distrito Federal | 3270.97 | 4139.05 | 2957.96 | 3261.49 | 3561.15 |
| Espírito Santo | - | - | - | - | - |
| Goiás | 4081.19 | 4889.6 | 4122.96 | 4362.55 | 9795.47 |
| Maranhão | 568.07 | 591.76 | 389.27 | 1031.92 | 606.9 |
| Mato Grosso | 973.33 | 1311.39 | 637.1 | 687.17 | 1638.31 |
| Mato Grosso do Sul | 964.76 | 870.86 | 713.08 | 3584.11 | 1414.12 |
| Minas Gerais | 27914.04 | 35223.9 | 19514.73 | 14264.82 | 53834.54 |
| Pará | 530.42 | 603.99 | 308.49 | 1133.26 | 1093.9 |
| Paraíba | 737.85 | 1031.91 | 275.54 | 568.89 | 543.3 |
| Paraná | 10187.53 | 14160.79 | 8957.75 | 5453.93 | 19491.39 |
| Pernambuco | 1219.49 | 1459.13 | 937.87 | 1626.65 | 1702.39 |
| Piauí | 339.3 | 317.46 | 321.3 | 190.65 | 502.38 |
| Rio de Janeiro | 4392.15 | 10707.1 | 3344.29 | 15883.43 | 7007.1 |
| Rio Grande do Norte | 504.33 | 662.92 | 416 | 305.82 | 402.79 |
| Rio Grande do Sul | 6101.44 | 7212.6 | 6512.75 | 2874.93 | 6786.81 |
| Rondônia | 271.28 | 320.94 | 209.99 | 513.51 | 362.63 |
| Roraima | 76.95 | 74.46 | 57.65 | 55.55 | 70.32 |
| Santa Catarina | 11577.1 | 11317.19 | 10251.62 | 8830.4 | 15341.5 |
| São Paulo | 59150.61 | 60836.59 | 47280 | 55802.34 | 102921.67 |
| Sergipe | 128.07 | 206.87 | 193.35 | 761.92 | 208.83 |
| Tocantins | 316.82 | 415.83 | 251.67 | 241.73 | 333.54 |

**Fig 4.**
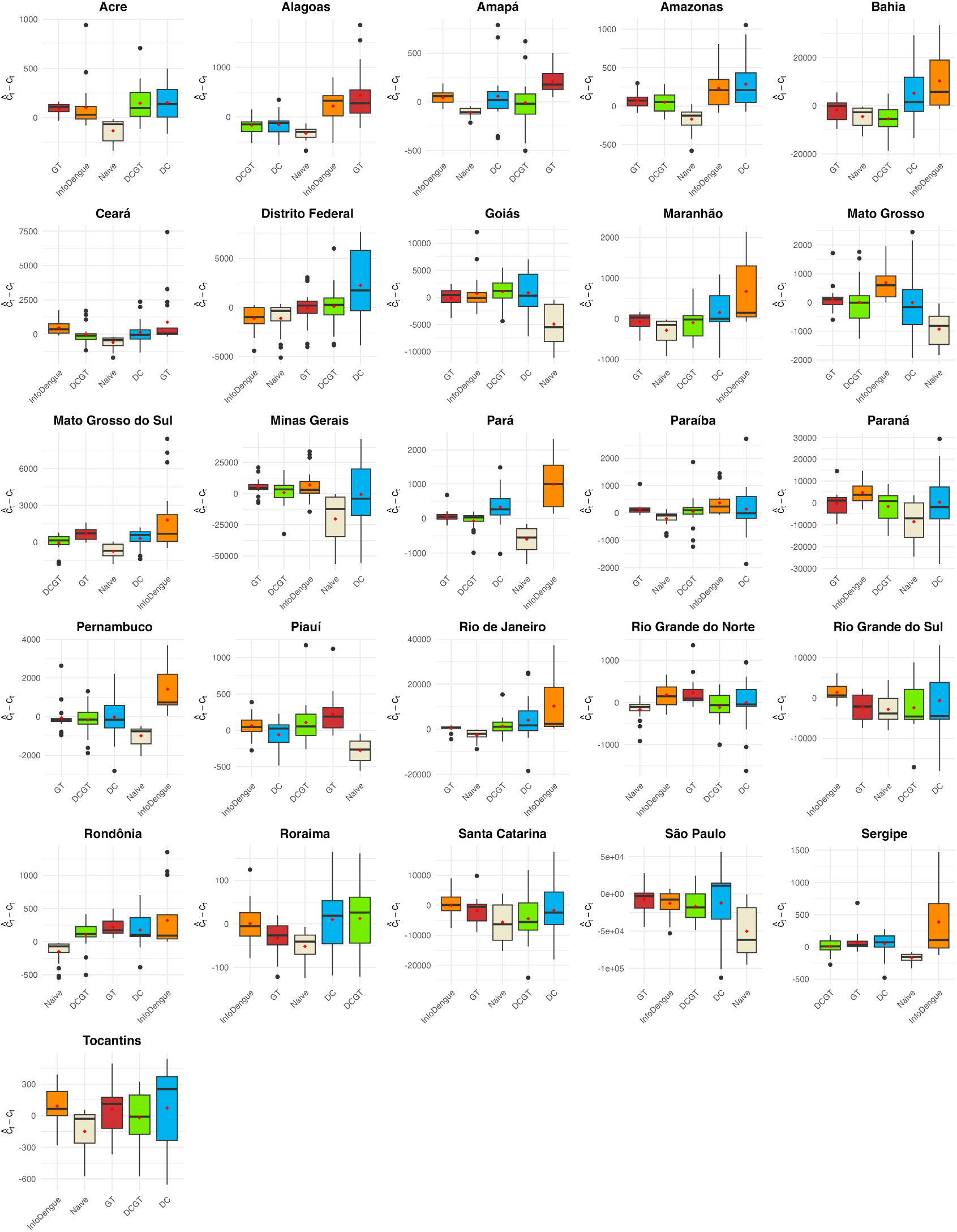
Boxplots of differences between the nowcasts and actual cases in Brazilian states. Models are sorted in ascending order of their mean absolute difference.

**Fig 5.**
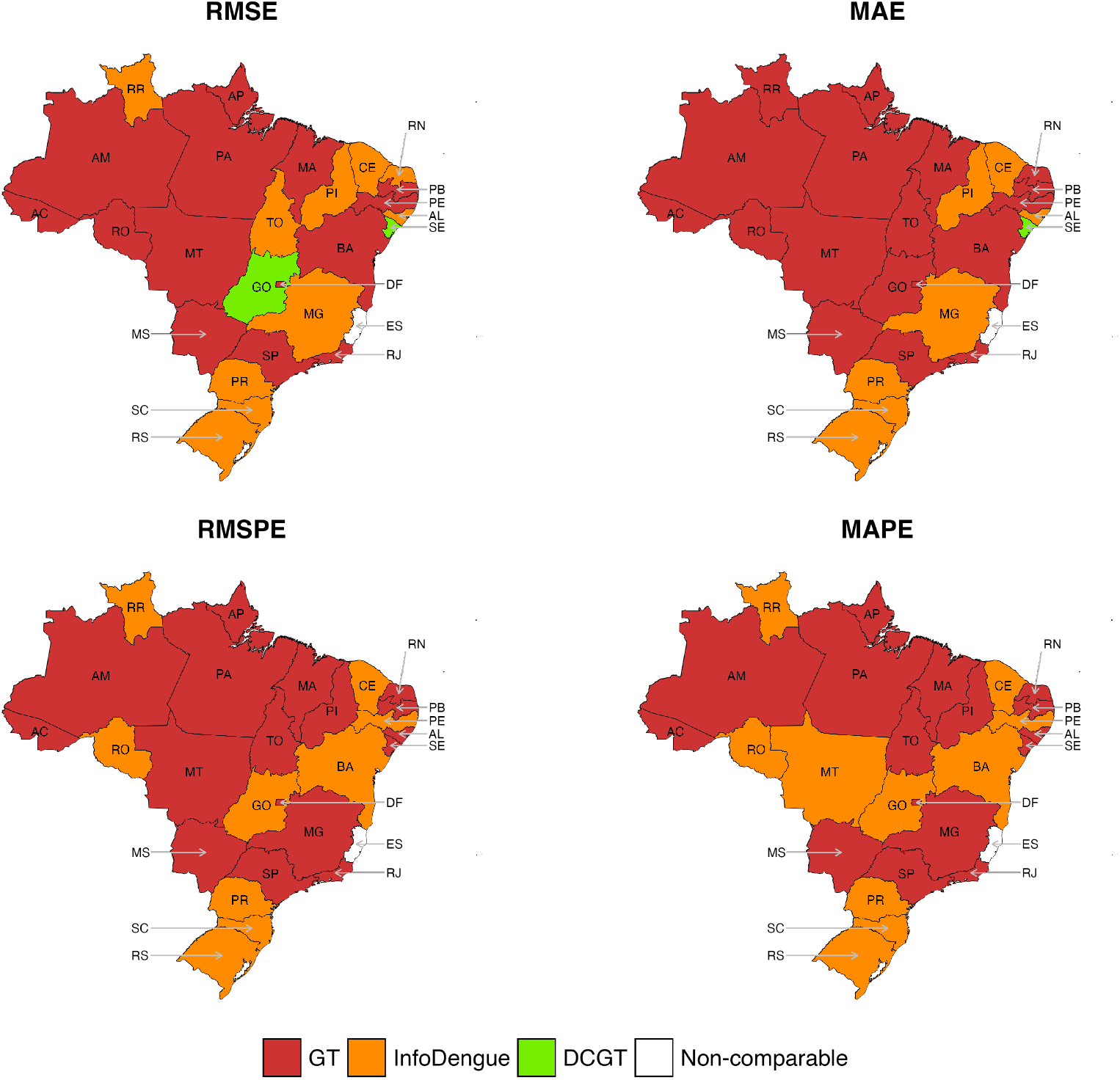
Best nowcasting models in different states according to different metrics.

Table 1 presents the RMSE values for prediction errors of each model. Overall, the GT model performed the best among the five approaches, achieving the most accurate predictions in 14 out of 26 states evaluated. The InfoDengue and DCGT models showed comparable performance, excelling in 10 and 2 states, respectively. Conversely, the naive and DC approaches performed the worst. In several states, such as Acre, Pará, Paraíba, Piauí, Rio Grande do Sul, and Sergipe, the GT model significantly outperformed the second-best models (the error of the second-best performing model was more than 1.5 times the error of the best-performing model). The smallest errors were observed in Roraima, all below 100, whereas São Paulo exhibited the largest errors, ranging from approximately 50,000 to 100,000, reflecting the differences in population sizes.

For MAE (Table 2), the overall results were similar to those observed for RMSE. The GT model outperformed the other approaches in 18 out of 26 states, followed by InfoDengue and DCGT, which were better in 7 and 1 states, respectively. The naive approach and the DC model both had the poorest performance, with no states showing an advantage. For Acre, Ceará, Maranhão, Pará, Paraíba, Paraná, and Rio Grande do Sul, the difference between the best and second-best performances was significant.

**Table 2.** MAE obtained for each state and nowcasting approach. Red and blue represent the best and the second best performances respectively (the lowest and the second lowest error).

|  | DCGT | DC | GT | InfoDengue | Naive |
| --- | --- | --- | --- | --- | --- |
| Acre | 188.31 | 211.99 | 85.65 | 142.43 | 217.21 |
| Alagoas | 338.76 | 374.89 | 471.43 | 259.64 | 491.14 |
| Amapá | 226.2 | 194.61 | 97.75 | 115.43 | 289.29 |
| Amazonas | 127.59 | 334.89 | 93.89 | 264 | 267 |
| Bahia | 9133.97 | 9803.59 | 5523.38 | 12433.25 | 7719 |
| Ceará | 713.36 | 775.02 | 1122.91 | 494.57 | 889.5 |
| Distrito Federal | 2419.85 | 3545.86 | 2110.1 | 2540.79 | 2553.86 |
| Espírito Santo | - | - | - | - | - |
| Goiás | 3474.26 | 4057.95 | 3436.14 | 3546.32 | 9081.64 |
| Maranhão | 507.33 | 477.82 | 279.01 | 790.43 | 498.86 |
| Mato Grosso | 841.84 | 1102.02 | 524.69 | 535.29 | 1565.5 |
| Mato Grosso do Sul | 681.83 | 735.97 | 604.91 | 2354.96 | 1294.5 |
| Minas Gerais | 20838.81 | 27981.08 | 14765.98 | 11173.39 | 46475.79 |
| Pará | 386.59 | 481.63 | 255.89 | 939.14 | 1011.86 |
| Paraíba | 479.25 | 735.24 | 177.42 | 385.86 | 393.86 |
| Paraná | 8195.76 | 12612 | 7558.59 | 4754.68 | 16972.29 |
| Pernambuco | 975.58 | 1147.88 | 769.57 | 1370.32 | 1585.29 |
| Piauí | 241.91 | 247.18 | 214.02 | 158.86 | 487.21 |
| Rio de Janeiro | 2882.21 | 8094.71 | 2302.94 | 11372.46 | 5840.71 |
| Rio Grande do Norte | 370.4 | 494.38 | 249.93 | 256.39 | 274.43 |
| Rio Grande do Sul | 5442.33 | 6910.77 | 5441.37 | 2456.43 | 6150 |
| Rondônia | 214.77 | 273.16 | 189.38 | 348.57 | 267.5 |
| Roraima | 61.58 | 61.47 | 41.89 | 45.89 | 57.71 |
| Santa Catarina | 9879.94 | 8960.95 | 8800.07 | 6889.75 | 13280.21 |
| São Paulo | 56458.59 | 45628.1 | 43625.64 | 49304.36 | 98125.07 |
| Sergipe | 103.47 | 173.06 | 108.79 | 552.18 | 196.14 |
| Tocantins | 251.01 | 364.25 | 202.12 | 208.32 | 243.43 |

RMSPE (Table 3) and MAPE (Table 4) values indicate that the top two models remain GT and InfoDengue, as they consistently produce the best nowcasts across all 26 states. For RMSPE, GT leads in 17 states, and InfoDengue in 11 states (both models produce the same RMSPE in Mato Grosso and Tocantins). Moreover, in many states, there is a notable difference between the performance of the best and second-best models. For example, in Alagoas, Amapá, Mato Grosso do Sul, and Rio Grande do Sul, the error of the second-best performing model was more than 2 times the error of the best-performing model. The results are similar for MAPE, with the GT model maintaining its lead in 16 states, followed by InfoDengue in 11 states (they are both the best in Acre).

**Table 3.** RMSPE obtained for each state and nowcasting approach. Red and blue represent the best and the second best performances respectively (the lowest and the second lowest error).

|  | DCGT | DC | GT | InfoDengue | Naive |
| --- | --- | --- | --- | --- | --- |
| Acre | 0.41 | 0.43 | 0.27 | 0.28 | 1.29 |
| Alagoas | 1.33 | 1.71 | 0.35 | 1.03 | 5.88 |
| Amapá | 2.94 | 4.34 | 0.29 | 3.35 | 71.04 |
| Amazonas | 0.21 | 0.35 | 0.16 | 0.25 | 0.62 |
| Bahia | 13.36 | 0.6 | 0.65 | 0.45 | 1.8 |
| Ceará | 0.44 | 0.63 | 0.33 | 0.23 | 0.96 |
| Distrito Federal | 0.31 | 1.84 | 0.26 | 0.43 | 0.42 |
| Espírito Santo | - | - | - | - | - |
| Goiás | 0.27 | 0.38 | 0.25 | 0.24 | 1.01 |
| Maranhão | 0.94 | 1.31 | 0.42 | 0.44 | 1.28 |
| Mato Grosso | 0.39 | 0.6 | 0.21 | 0.21 | 1.23 |
| Mato Grosso do Sul | 0.67 | 0.41 | 0.19 | 0.58 | 4.42 |
| Minas Gerais | 0.37 | 2.78 | 0.24 | 0.79 | 2.1 |
| Pará | 0.46 | 0.47 | 0.21 | 0.34 | 1.92 |
| Paraíba | 0.56 | 1.88 | 0.15 | 0.23 | 0.49 |
| Paraná | 0.56 | 0.37 | 0.21 | 0.14 | 0.63 |
| Pernambuco | 0.63 | 1.49 | 0.32 | 0.29 | 1.02 |
| Piauí | 0.33 | 0.79 | 0.23 | 0.58 | 1.57 |
| Rio de Janeiro | 0.86 | 0.71 | 0.22 | 0.4 | 0.9 |
| Rio Grande do Norte | 0.45 | 1.25 | 0.16 | 0.19 | 0.34 |
| Rio Grande do Sul | 2.01 | 0.63 | 0.59 | 0.21 | 0.64 |
| Rondônia | 0.53 | 0.52 | 0.44 | 0.36 | 1.63 |
| Roraima | 0.82 | 0.78 | 0.72 | 0.45 | 1.3 |
| Santa Catarina | 2.15 | 0.57 | 0.43 | 0.35 | 0.85 |
| São Paulo | 0.56 | 1.31 | 0.35 | 0.45 | 1.32 |
| Sergipe | 0.3 | 0.73 | 0.2 | 0.48 | 0.93 |
| Tocantins | 0.36 | 0.46 | 0.23 | 0.23 | 0.44 |

**Table 4.** MAPE obtained for each state and nowcasting approach. Red and blue represent the best and the second best performances respectively (the lowest and the second lowest error).

|  | DCGT | DC | GT | InfoDengue | Naive |
| --- | --- | --- | --- | --- | --- |
| Acre | 0.34 | 0.39 | 0.22 | 0.22 | 1.07 |
| Alagoas | 0.94 | 1.18 | 0.3 | 0.55 | 2.73 |
| Amapá | 1.88 | 2.05 | 0.24 | 1.53 | 40.38 |
| Amazonas | 0.18 | 0.31 | 0.13 | 0.23 | 0.52 |
| Bahia | 5.26 | 0.48 | 0.5 | 0.39 | 1.12 |
| Ceará | 0.38 | 0.43 | 0.22 | 0.18 | 0.86 |
| Distrito Federal | 0.27 | 1.04 | 0.22 | 0.4 | 0.37 |
| Espírito Santo | - | - | - | - | - |
| Goiás | 0.22 | 0.28 | 0.22 | 0.2 | 0.96 |
| Maranhão | 0.78 | 0.82 | 0.34 | 0.37 | 1.13 |
| Mato Grosso | 0.34 | 0.48 | 0.18 | 0.16 | 1.17 |
| Mato Grosso do Sul | 0.41 | 0.32 | 0.18 | 0.46 | 2.21 |
| Minas Gerais | 0.3 | 1.46 | 0.2 | 0.42 | 1.77 |
| Pará | 0.3 | 0.31 | 0.17 | 0.31 | 1.76 |
| Paraíba | 0.36 | 1.16 | 0.12 | 0.19 | 0.39 |
| Paraná | 0.31 | 0.31 | 0.18 | 0.12 | 0.56 |
| Pernambuco | 0.46 | 0.88 | 0.29 | 0.27 | 0.98 |
| Piauí | 0.28 | 0.55 | 0.19 | 0.31 | 1.45 |
| Rio de Janeiro | 0.46 | 0.57 | 0.18 | 0.35 | 0.83 |
| Rio Grande do Norte | 0.33 | 0.7 | 0.13 | 0.17 | 0.25 |
| Rio Grande do Sul | 0.99 | 0.57 | 0.51 | 0.18 | 0.6 |
| Rondônia | 0.47 | 0.5 | 0.39 | 0.34 | 1.47 |
| Roraima | 0.62 | 0.64 | 0.53 | 0.39 | 1.05 |
| Santa Catarina | 1 | 0.43 | 0.38 | 0.28 | 0.75 |
| São Paulo | 0.51 | 0.66 | 0.33 | 0.39 | 1.27 |
| Sergipe | 0.23 | 0.45 | 0.16 | 0.44 | 0.84 |
| Tocantins | 0.29 | 0.4 | 0.19 | 0.2 | 0.31 |

Table 5 displays the 95% coverage probabilities, and Table 6 the average width of 95% uncertainty intervals obtained for each state and nowcasting approach. In these tables, results for InfoDengue are not included, as nowcasts for this method are provided by municipality level, while our analysis is conducted at the state level. We found that both the DC and GT models exhibited high coverage rates, achieving the highest coverage in 14 different states. In contrast, the DCGT model only achieved the highest coverage in 6 states. While the coverage rates for DC and GT are comparable, the average width of the uncertainty intervals in Table 6 shows that the DC model has a significantly larger average width. In some regions, such as the Distrito Federal and Maranhão, the prediction width for DC can be as much as twice that of GT.

**Table 5.** 95% coverage probabilities obtained for each state and nowcasting approach. Red and blue represent the best and the second best performances respectively (the highest and the second highest coverage probabilities).

|  | DCGT | DC | GT |
| --- | --- | --- | --- |
| Acre | 1 | 1 | 1 |
| Alagoas | 0.79 | 0.79 | 0.71 |
| Amapá | 0.5 | 0.71 | 0.93 |
| Amazonas | 1 | 0.86 | 1 |
| Bahia | 0.14 | 0.29 | 0.29 |
| Ceará | 0.64 | 0.79 | 0.71 |
| Distrito Federal | 0.36 | 0.64 | 0.36 |
| Espírito Santo | - | - | - |
| Goiás | 0.43 | 0.64 | 0.36 |
| Maranhão | 0.21 | 0.43 | 0.57 |
| Mato Grosso | 0.43 | 0.43 | 0.79 |
| Mato Grosso do Sul | 0.79 | 1 | 0.79 |
| Minas Gerais | 0.43 | 0.57 | 0.43 |
| Pará | 0.5 | 0.64 | 0.5 |
| Paraíba | 0.71 | 0.64 | 0.93 |
| Paraná | 0.43 | 0.36 | 0.36 |
| Pernambuco | 0.57 | 0.64 | 0.71 |
| Piauí | 0.71 | 0.93 | 0.79 |
| Rio de Janeiro | 0.79 | 0.64 | 0.71 |
| Rio Grande do Norte | 0.71 | 0.86 | 0.86 |
| Rio Grande do Sul | 0.29 | 0.21 | 0.29 |
| Rondônia | 0.86 | 0.86 | 1 |
| Roraima | 0.46 | 0.57 | 0.79 |
| Santa Catarina | 0.29 | 0.36 | 0.21 |
| São Paulo | 0.07 | 0.43 | 0.14 |
| Sergipe | 0.86 | 0.71 | 0.93 |
| Tocantins | 0.93 | 1 | 1 |

**Table 6.** Average width of 95% uncertainty intervals obtained for each state and nowcasting approach. Red and blue represent the best and the second best performances respectively (the smallest and the second smallest interval width).

|  | DCGT | DC | GT |
| --- | --- | --- | --- |
| Acre | 1369.75 | 1496.42 | 934.25 |
| Alagoas | 940.72 | 1078.19 | 999.65 |
| Amapá | 357.29 | 396.01 | 350.97 |
| Amazonas | 673.59 | 1183.24 | 611.41 |
| Bahia | 4701.96 | 9283.53 | 4196.98 |
| Ceará | 2282.71 | 2590.75 | 2768.77 |
| Distrito Federal | 2904.71 | 10230.87 | 2930 |
| Espírito Santo | - | - | - |
| Goiás | 6422.55 | 9791.6 | 4802.93 |
| Maranhão | 414.36 | 695.88 | 326.71 |
| Mato Grosso | 1661.14 | 1760.56 | 1533.96 |
| Mato Grosso do Sul | 2893.6 | 3580.1 | 2048.75 |
| Minas Gerais | 17764.49 | 44231.53 | 15694.19 |
| Pará | 498.1 | 1035.91 | 491.75 |
| Paraíba | 873.72 | 1367.52 | 785.07 |
| Paraná | 11586.03 | 20746.87 | 9641.61 |
| Pernambuco | 1640.81 | 2390.35 | 1880.34 |
| Piauí | 512.8 | 1202.53 | 492.37 |
| Rio de Janeiro | 6039.16 | 12327.16 | 4817.5 |
| Rio Grande do Norte | 972.67 | 1587.1 | 822.89 |
| Rio Grande do Sul | 5405.35 | 6932.19 | 3999.01 |
| Rondônia | 887.8 | 1032.86 | 654.64 |
| Roraima | 124.69 | 128.45 | 138.4 |
| Santa Catarina | 10271.47 | 12404.89 | 5301.92 |
| São Paulo | 31365.06 | 52015.13 | 28757.8 |
| Sergipe | 328.61 | 393.69 | 356.98 |
| Tocantins | 1161.9 | 1624.41 | 1215.73 |

Additionally, there is considerable variation in 95% coverage probabilities across states. For example, in Acre, Amazonas, and Tocantins, all models have high coverage rates, approaching or reaching 1. However, in states like Bahia and Rio Grande do Sul, the coverage rates for all models are only around 20% to 30%. The variation also exists in average width. This variation may be influenced by differences in population size, data quality, or the nature of dengue transmission across states, leading to significant discrepancies in results.

Figure 6 shows the weekly nowcasting results for each method. The green lines represent the reported cases for a given epidemiological week after 10 weeks. We consider these values as the true number of cases to benchmark the models’ performance. Notably, the suspected cases lines are consistently the lowest, serving as a practical “lower bound”, with various models employed to adjust this lower limit. In general, the red lines representing nowcasts obtained with Google Trends closely align with the true values, indicating that the GT model performs particularly well. In contrast, the purple line for InfoDengue nowcasts frequently appear above the true values, suggesting a tendency to overestimate potential cases. Despite this, the InfoDengue nowcasts successfully capture the trends indicated by the true values. Finally, the DCGT and GT nowcasts are generally less likely to accurately capture the trends of the true values.

**Fig 6.**
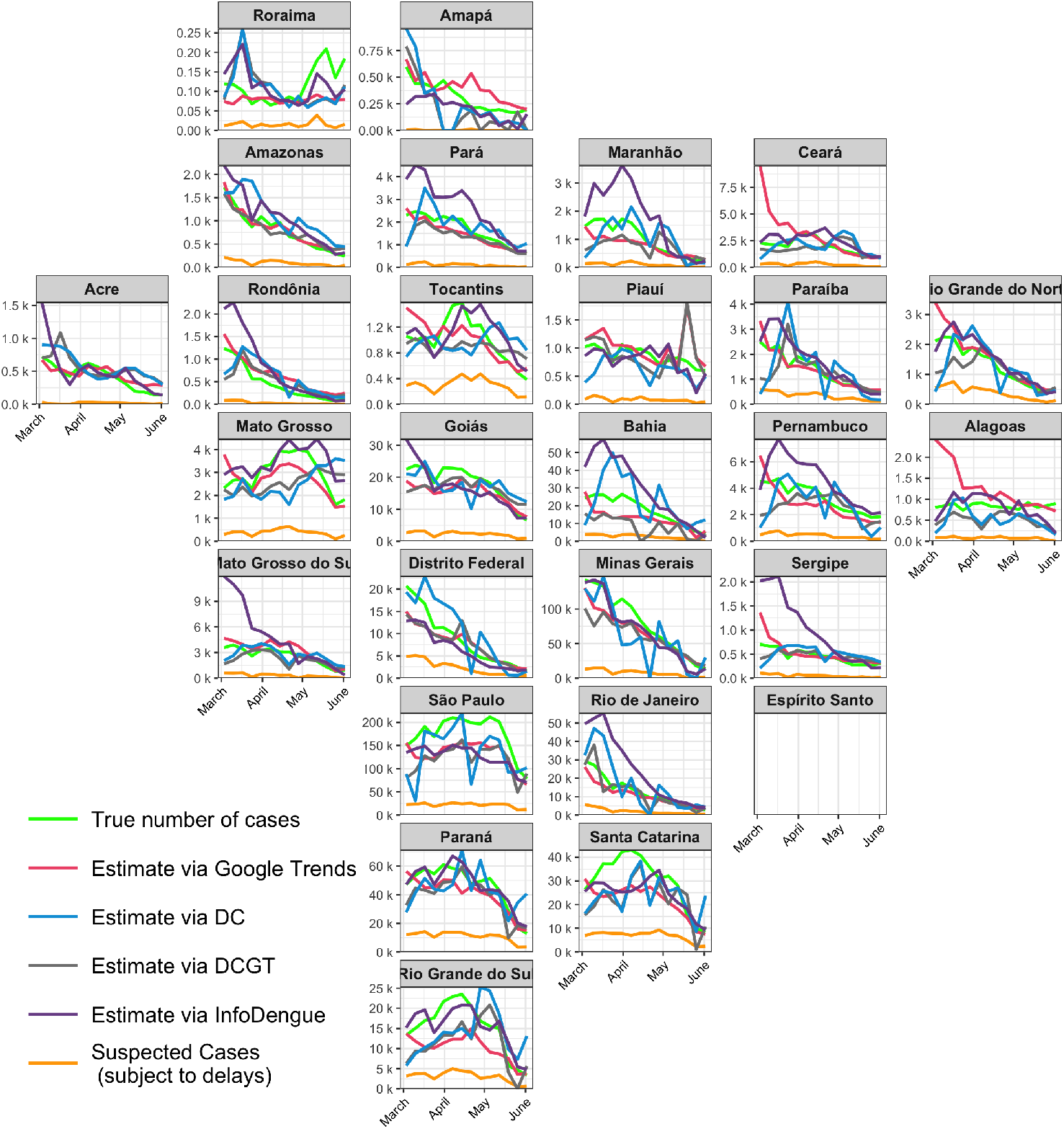
Predictions of the models used in the study from week 10 to week 23 of 2024 (March 3 to June 2, 2024). The ”True number of cases” line represents the reported cases after 10 weeks, which are used to evaluate the performance of the models.

### Dengue Tracker website

Timely and accurate information on dengue cases is crucial for prevention and control. We developed the Dengue Tracker website (https://diseasesurveillance.github.io/dengue-tracker/) to provide weekly updates on the number of official dengue cases per state in Brazil, as well as at the country level. Additionally, the website provides corrected case counts incorporating information from Google Trends (using our GT model) and also the InfoDengue results. We believe these reports will assist policymakers in understanding dengue levels and guide their decisions.

Each week, the number of dengue cases is downloaded through InfoDengue’s API, and the Google Trends information for the specific keywords is downloaded from https://trends.google.com/trends/. Data are download from the last 5 years up to the week we are interested in nowcasting. At the country level, Dengue Tracker shows the dengue incidence rate through an interactive choropleth map. Besides, it depicts the time series of the number of cases, the fitted model, and the corrections from both our model and InfoDengue’s for each state in a plot with the shape of Brazil. Reports are also provided for each state. The website is built using RMarkdown (34) and GitHub Pages. The graphic components are built using ggplot2 (35), plotly (36), geofacet (37) and leaflet (38).

## Discussion

Reporting delays in surveillance data make traditional surveillance systems ineffective for planning and control. In this paper, we compared the usefulness of integrating Google Trends information for dengue nowcasting in Brazil with other analytical approaches that rely solely on reported data. Specifically, we evaluated the error and uncertainty produced by approaches that used only Google Trends information (GT), only dengue case data (DC), and a combination of both (DCGT). In addition, we compared these results with the nowcasting algorithm provided in InfoDengue and a naive approach, where the number of cases in a given week was nowcasted as the number of cases reported in the previous week.

Our study demonstrates the effectiveness of combining Google Trends information with reported case data for nowcasting dengue in Brazil. We show that using the reported number of cases as the nowcast for the following week, as done in the naive approach, is insufficient for real-time monitoring since it significantly underestimates the actual number of dengue cases. Overall, the GT model demonstrates the lowest error across most states, outperforming other models in all metrics, with InfoDengue ranking second. In contrast, neither DCGT nor DC provides significant improvements in nowcasting accuracy. The results, as illustrated through boxplots and accuracy metrics, confirm that incorporating Google Trends data effectively enhances predictive performance. Regarding uncertainty, we lack uncertainty intervals for InfoDengue since the nowcasts are provided at the municipality level. Among the remaining models, GT generally has the narrowest intervals while achieving the highest coverage rate. In contrast, DCGT generates significantly narrower uncertainty intervals compared to DC but slightly lowers the coverage rate. Overall, GT improves predictive accuracy and reduces uncertainty.

In our study, we excluded Espírito Santo from model comparisons because the state ceased reporting dengue cases. However, it is possible to fit an appropriate GT model using historical data and continue nowcasting with Google Trends data, even after reporting has stopped — this approach was actually implemented on the Dengue Tracker website. Another noteworthy case is Rio de Janeiro, where changes in the notification system’s infrastructure and workflow were introduced during the epidemic to achieve a faster response. This shift was not captured by InfoDengue, leading to nowcasts that significantly overestimated the actual number of cases from March to April, whereas GT’s nowcasts were much closer to the true figures.

One limitation of our study is that Google Trends data is inherently biased since not all individuals use Google to search for dengue-related information. This bias may result in underrepresentation of specific populations or regions, potentially affecting the accuracy of our models. Further research is needed to understand how different population groups use Google to search for dengue information, how to select dengue-related keywords that accurately reflect disease transmission, and to develop models that integrate Google Trends and dengue case data in the most effective way.

In this study, we employed statistical models that excluded the most recent weeks of incomplete information to generate dengue nowcasts. Although this approach allowed us to demonstrate the superior performance of approaches using Google Trends data compared to models relying only on reported cases, further work could be done to develop models that utilize incomplete data to further improve predictive accuracy.

In addition, the models used in this study are limited in their ability to detect sudden changes in dengue incidence, since they heavily rely on historical data. This limitation hinders their effectiveness in identifying abrupt outbreaks or sharp increases in cases. Future research will explore more flexible approaches to improve responsiveness.

Additionally, we will investigate the incorporation of variables like climate and socio-economic factors, known to influence dengue transmission, into future models. Moreover, we intend to develop spatial models that allow us to obtain nowcasts at finer geographical resolutions, such as microgregion or municipality levels (39). This enhancement will provide localized nowcasts, enabling more precise public health interventions.

Effective and timely communication of dengue activity levels is crucial for planning and response efforts in public health. To address this need, we also developed Dengue Tracker (https://diseasesurveillance.github.io/dengue-tracker/index.html), a system designed to aggregate, analyze, and visualize dengue case data. This system provides weekly nowcasts at the state level in Brazil, aiding decision-makers and the public in understanding current risk levels. The Dengue Tracker website is updated weekly and features interactive maps and time series plots that dynamically present the latest dengue information across Brazil. By integrating our nowcasting models that use Google Trends information into this platform, the website delivers real-time alerts and trend analyses for better disease prevention and control.

In conclusion, our study presents a promising approach to improving dengue surveillance in Brazil, emphasizing the potential of integrating digital data with traditional epidemiological models. This integration significantly enhances situational awareness for public health authorities and the general public alike, facilitating responses to changes in dengue activity levels, ultimately reducing the impact of dengue and improving the health and well-being of the population.

## Data Availability

For reproducibility purposes, data and code to apply the methods presented in the manuscript are provided in the GitHub repository: https://github.com/diseasesurveillance/dengue-tracker/tree/main/paper

## Acknowledgements

This research received financial support from The Letten Prize (https://lettenprize.com/), with a personal award to Paula Moraga. Rafael Izbicki is grateful for the financial support of CNPq (422705/2021-7 and 305065/2023-8) and FAPESP (grant 2023/07068-1).

